# PDIVAS: Pathogenicity predictor for Deep-Intronic Variants causing Aberrant Splicing

**DOI:** 10.1101/2023.03.20.23287464

**Authors:** Ryo Kurosawa, Kei Iida, Masahiko Ajiro, Tomonari Awaya, Mamiko Yamada, Kenjiro Kosaki, Masatoshi Hagiwara

**Author notes:** Correspondence should be addressed to R.K. and M.H., This work was done in Kyoto, Japan. Corresponding author’s address: Yoshida-Konoe-cho, Sakyo-ku, Kyoto, 606-8501, JAPAN.

## Abstract

Deep-intronic variants often cause genetic diseases by altering RNA splicing. However, these pathogenic variants are overlooked in whole-genome sequencing analyses, because they are quite difficult to segregate from a vast number of benign variants (approximately 1,500,000 deep-intronic variants per individual). Therefore, we developed the Pathogenicity predictor for Deep-Intronic Variants causing Aberrant Splicing (PDIVAS), an ensemble machine-learning model combining multiple splicing features and regional splicing constraint metrics. Using PDIVAS, around 27 pathogenic candidates were identified per individual with 95% sensitivity, and causative variants were more efficiently prioritized than previous predictors in simulated patient genome sequences. PDIVAS is available at https://github.com/shiro-kur/PDIVAS.

## Background

Causative variants of Mendelian diseases remain to be determined in 50-75% of patients, regardless of the technical progress of whole-exome sequencing (WES) and whole-genome sequencing (WGS) [1-4]. One of the major obstacles in the diagnostic process is the technical difficulty of evaluating genetic variants in deep-intronic regions. Intronic variants with splice alterations have been reported as the causative variants of dystrophinopathy, neurofibromatosis type I, and inherited retinal diseases [5-10] because these splice-altering variants (SAVs) create pathogenic pseudoexons or extend existing exons, by affecting recognition by splicing factors (e.g., small nuclear ribonucleoprotein and RNA-binding proteins). (Fig. 1a). The resulting splicing alterations subsequently lead to mRNA destabilization by nonsense-mediated decay (NMD) or functional defects in encoded proteins. Previously, most of the pathogenic deep-intronic SAVs were discovered through the “RNA-based” diagnosis using RT-PCR or RNA sequencing on patient-derived cells or tissues [11-13]. However, causative genes are often expressed in patients’ specific tissues, such as the brain and heart, which are rarely available to clinicians.

**Fig. 1.**
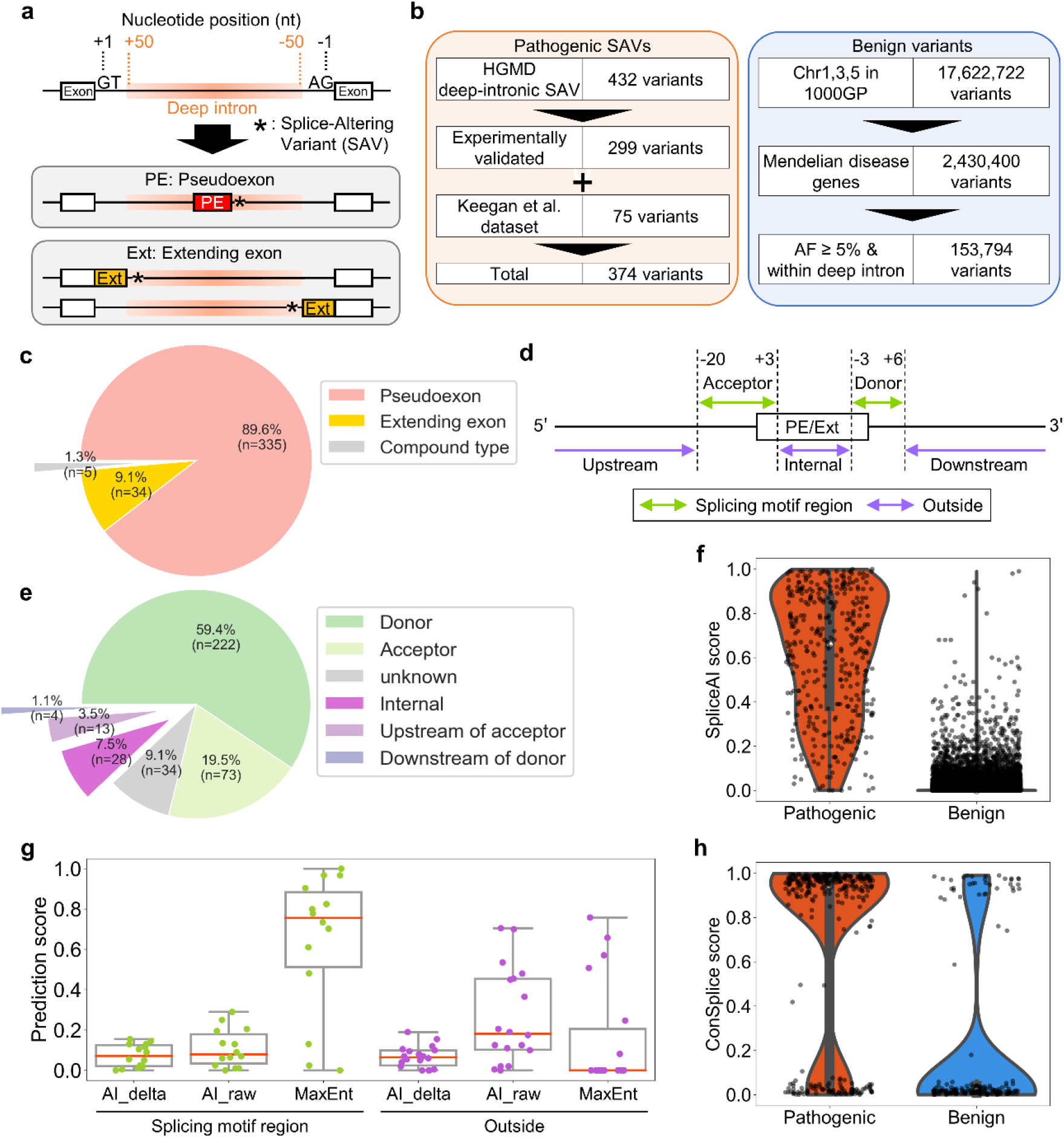
Prospective features to classify pathogenic and benign variants in a curated dataset. **a** Schematic representation of aberrant splicing induced by deep-intronic splice-altering variants (SAVs). **b** The curated dataset comprised pathogenic deep-intronic SAVs from the Human Gene Mutation Database (HGMD) and pseudo exon dataset by Keegan et al. [7] and benign deep-intronic variants from the 1000 Genomes Project (1000GP). **c** Splice-type classification of pathogenic SAVs in the curated dataset. Compound type refers to the SAVs that cause both a pseudoexon and an extending exon in the intron region. **d** Definition of region names based on the relative position of the splice sites of pseudoexon (PE) or extending exon (Ext). **e** Classification of pathogenic SAVs on their relative positions to the nearest splice sites of the pseudoexon or extending exons. **f** Violin plot, box plot, and strip plot depicting maximums of SpliceAI donor/acceptor delta gain scores for variants in the curated dataset. **g** Violin plot, box plot, and strip plot depicting prediction scores by mean values of SpliceAI donor/acceptor delta gain scores, mean values of SpliceAI donor/acceptor raw gain scores, and MaxEntScan scores for pathogenic SAVs with ≤ 0.2 maximums of SpliceAI donor/acceptor delta gain scores (n=44). **h** Violin plot and strip plot depicting ConSplice scores (human splicing constraint metrics) for each variant in the curated dataset with > 0.2 maximums of SpliceAI donor/acceptor delta gain scores (n=190). AF, Allele frequency

In contrast to the RNA samples, the genetic variants of almost all genes can be identified through WGS analysis on blood or skin samples, which are readily accessible. However, distinguishing pathogenic deep-intronic variants from the vast number of benign variants is challenging owing to the presence of over 1,800,000 intronic variants in an individual [14]. Therefore, the variants must be filtered through a computational process with pathogenicity predictors to enable clinicians to manually evaluate the candidate variants.

In this study, we present a novel pathogenicity predictor for deep-intronic variant prioritization, called Pathogenicity predictor for Deep-Intronic Variant causing Aberrant Splicing (PDIVAS). We demonstrate that PDIVAS offers a clinically applicable strategy for deep-intronic variant prioritization in the diagnosis of rare genetic diseases.

## Method

### Variant annotation

The effect of variants on genes was annotated by the ensemble variant effect predictor (VEP) (version 105, GRCh37) by referring to one transcript per genome region selected by the “--pick_allele_gene” option. The transcript annotation was based on GENCODE V19 [15, 16]. We selected deep-intronic variants according to HGVS nomenclature that describes the relative position of the variant within the gene [17]. The annotation of allele frequency in the gnomAD WGS database (r.2.0.1) was conducted through the VEP “--custom” option referring to the tabix-indexed VCF file downloaded from the Ensembl FTP site [18].

The features from SpliceAI (delta_gain_max, delta_gain_mean, and raw_gain_mean) were obtained from the output-customized SpliceAI whose original version was 1.3.1. By default, SpliceAI only outputs four delta scores of acceptor gain/loss and donor gain/loss, which are calculated by subtracting the splice site score on reference from the alternative sequence with a variant of interest [19]. Our customized SpliceAI script outputs the raw scores for the alternative sequence before subtraction, as well. Furthermore, while default SpliceAI converts the delta scores: acceptor/donor gain to zero when the predicted site matches the nearest annotated splice site on the gene (with the “-m” mask option), we extended the mask function to all of the referred annotated splice sites on the gene to reduce the number of false positives. The SpliceAI delta_gain_max and mean scores were calculated as the maximum and mean values of acceptor and donor gain delta scores, respectively. The SpliceAI raw_gain_mean score was calculated as the mean value of the raw scores of acceptors and donors. And finally, SpliceAI predictions were performed with the distance option of “-d 300”.

The ConSplice feature was obtained from the score-precomputed bed file of the best_splicing_constraint_model provided by the authors [20]. MaxEntScan prediction of the variant’s effect on splicing was performed using the plugin module of VEP [21-23]. The feature-extraction algorithm from MaxEntScan is described in Supplemental Fig. 1, which corresponds to the interpretation algorithm of Shamsani et al. [23].

Predictors to be compared to PDIVAS were SpliceAI, Pangolin [24], ConSpliceML [20], MaxEntScan, and CADD-Splice [25], which were selected based on the following criteria: 1) the program or the precomputed score file is freely available, 2) the program can assess deep-intronic variants, and 3) the program is operated in a Linux environment and can be applied to large-scale variant analysis. SQUIRLS and SPiP also matched these criteria, but their developers recognized their lower performance on deep-intronic SAVs because of the limited number of training datasets [26, 27]. Therefore, we did not include them in this comparison. Pangolin (v1.0.1) was installed and run with the mask option (--mask True) and distance option (-d 300). To annotate ConSpliceML, we downloaded the score-precomputed VCF file provided by the authors and converted the genome version from GRCh38 to GRCh37 using Picard LiftoverVcf (v.2.27.1). The annotation of ConSpliceML was conducted through the VEP “--custom” option, referring to the precomputed VCF file on the GRCh37 version. Annotations in CADD-Splice (v1.6) were conducted on plugin modules of the Variant Effect Predictor (VEP). BCFtools, bash scripts, cyvcf2, and Python scripts were used to process the VCF files used in this research [28, 29].

### Curated dataset of deep-intronic variants

To focus solely on the variants within genes responsible for Mendelian diseases, we collected gene lists from the Online Mendelian Inheritance in Man (OMIM) and the Clinical Genomic Database (CGD) [30, 31]. In the OMIM gene list, there were many genes whose causative relationships with the registered phenotype were unclear, as well as those that only contributed to the susceptibility to multifactorial diseases (e.g., diabetes and asthma). To only focus on the genes whose contribution to the phenotype is clear in a way of Mendelian inheritance, we extracted genes with annotations of the mode of inheritance (autosomal dominant/recessive, X-linked, Y-linked), and the phenotype mapping key of the molecular basis of the diseases is known. Then, the OMIM and CGD gene lists were combined and non-coding genes were filtered out according to the GENCODE V19 annotation, resulting in a final list of Mendelian disease genes (https://github.com/shiro-kur/PDIVAS).

For the benign dataset, the variant lists from the 1000 Genomes Project (Phase 3, GRCh37) were downloaded from the UCSC FTP site. Variants on chromosomes 1, 3, and 5 whose sequences were not used to train SpliceAI were extracted to reduce data size. Additionally, copy number variants and variants in the multi-nucleotide variant format were removed because they could not be evaluated with many of the predictors used in this research. We annotated the remaining variants with VEP and extracted only deep intronic variants (≥50 bp away from the nearest splice sites) with ≥5% allele frequency in the 1000 Genomes Project and gnomAD population located on Mendelian disease genes. Subsequently, Pathogenic deep-intronic SAVs were collected from Human Gene Mutation Database Professional online (HGMD) 2020 and the Keegan et al. dataset [7, 32]. By utilizing the search function of HGMD, we extracted single nucleotide variants (SNVs) labeled with “disease-causing” and “splicing”, and located more than 30 bp away from the nearest splice sites. After we annotated the variants with VEP, only deep-intronic variants and variants with <1% allele frequency in the gnomAD population were extracted. The extracted variants were checked to determine if splice alterations were experimentally validated by reading all of the original reports. The variants whose splice alterations were validated using RT-PCR or RNA sequencing for patient RNA or minigene/midigene-expressed RNA were included in the final variant dataset. Other variants whose aberrant splicing was only predicted or not specified were filtered out. Keegan et al. [7] also cataloged deep-intronic SAVs with experimental validation in a process independent of HGMD and our reinspection. From the Keegan et al dataset, SNVs and short deletions (∼54nt) were obtained and annotated with VEP to extract only deep-intronic variants, those with <1% allele frequency in the gnomAD population, and located on the Mendelian disease genes. Non-overlapped SAVs were added to the reinforced HGMD dataset to complete the pathogenic dataset. Through the curation process, the relative variant positions to the nearest splice sites of the pseudoexons or extending exons were also checked and classified as “splicing motif region” or “outside” as described in Fig. 1d. The splicing motif region (region of the splice donor and acceptor site motif) conforms to the region defined in MaxEntScan [21].

### Model training and testing

For model training and evaluation of PDIVAS, a random forest model provided by scikit-learn (version 1.0.2) was used. We randomly split the curated dataset into 70% for model training and the remaining 30% for testing the trained model. Furthermore, within the training dataset, five-fold cross-validation was performed. In this process, the training dataset was equally split into five sub-datasets. Four of the five sub-datasets were used for parameter tuning of random forest, and the remaining sub-dataset was used to evaluate the trained model. This process was iteratively repeated for all five combinations of the training and test sub-datasets. Through five-fold cross-validation, the best combination of hyperparameters was determined when the average precision metrics, calculated as the mean of the five iteration results were maximized. The tuned hyperparameters in this model were: 1) the number of decision trees in the forest and the maximum depth of each tree; 2) the ratio of samples extracted from the whole training dataset to train each tree; and 3) the number of features to be used in each tree. Using these optimal hyperparameters, the five-fold cross-validation was conducted again to compare the classification performance of the intermediate models of PDIVAS with SpliceAI and Pangolin and evaluate the stability of the PDIVAS performance. Finally, the random forest model was trained on the entire training dataset using the optimized hyperparameters. The final PDIVAS model was evaluated on the test dataset with precision, recall, and Matthews correlation coefficient (MCC) metrics for each score threshold. To calculate the average precision, we computed the weighted mean of the precision achieved at each threshold, divided by the difference in recall from the previous threshold. This metric is more accurate than the area under the curve and does not require empirical curve construction [33]. We employed the average precision and MCC for the performance comparison because they were stable even when the classes in a dataset were highly imbalanced. The predictive performance of PDIVAS was compared to the SpliceAI_delta_gain_max, Pangolin gain score, ConSpliceML, MaxEntScan, and CADD-Splice Phred scores. ConSpliceML and CADD-Splice could not score some of the variants because their score-precomputed files did not cover all variant types, such as genomic insertion and deletion. When we calculated the statistical measures, we only referred to the variants with scores.

### Variant prioritization for the 1000 Genomes Project

We referred to the same variant lists of the 1000 Genomes Project under the sub-heading “Curated dataset of deep-intronic variants”, above. Variants on chrY were filtered out because the ConSplice score to calculate the PDIVAS score was not available there. Additionally, copy number variants and variants in the multi-nucleotide variant format were removed because they could not be evaluated with many of the predictors used in this study. From the remaining variants, deep-intronic variants of 20,731 protein-coding genes defined on GENCODE V19 were extracted. Subsequently, variants on Mendelian disease genes and variants with <1% allele frequency in both the 1000 Genomes Project and gnomAD populations were extracted as candidates for pathogenic variants. Finally, the rare variants were scored by PDIVAS, SpliceAI, and Pangolin and filtered by thresholds based on the sensitivity of pathogenic SAVs.

## Results

We developed PDIVAS, a novel machine-learning framework to evaluate the pathogenicity of deep-intronic variants. PDIVAS was constructed and optimized through (1) setting up a labeled dataset of known pathogenic and benign deep-intronic variants, (2) selecting a set of features to classify pathogenic and benign variants, and (3) training the random forest model with selected features. Finally, we evaluated the performance of PDIVAS versus previous predictors through the classification of pathogenic and benign deep-intronic variants, WGS analysis in control individuals, and simulation analysis of patient genome sequences.

### Curating a dataset consisting of pathogenic and benign deep-intronic variants

To train the PDIVAS model on truly pathogenic deep-intronic variants, we surveyed the original reports of the variants and collected only those pathogenic variants with experimentally validated splice alterations. As data sources of pathogenic deep-intronic SAVs, we referred to HGMD and a pathogenic pseudoexon dataset constructed by Keegan et al [7, 32]. From HGMD, we extracted 432 variants located within deep introns out of 26,610 SNVs annotated as both “disease-causing” and “splicing mutation” (Fig. 1b). In this study, we defined deep introns as intronic regions ≥50 bases from the nearest annotated splice sites (Fig. 1a). Reinspection of all the originally reported papers revealed that some lacked experimental validation of their occurrence of aberrant splicing by either splicing minigene/midigene assays or patient-derived RNA sample analysis. To construct a high-quality training dataset, we used only pathogenic SAVs with such experimental evidence. Furthermore, we focused on the Mendelian diseases genes and excluded polygenic diseases and those whose mode of inheritance was obscure. The lists of 4,429 genes (hereinafter called Mendelian disease genes) were constructed from the OMIM and the CGD. This filtering also clarifies the causality of the variants and the patient phenotypes. We then extracted variants with <1% allele frequency in the gnomAD population. These procedures yielded 290 variants curated as pathogenic SAVs from HGMD (Fig. 1b).

The SAVs curated by Keegan et al (n=359) were already checked with experimental validation. We also extracted those on Mendelian disease genes with <1% allele frequency in the gnomAD population. The obtained variants were added exclusively to the variant list obtained from HGMD. Our combined approach yielded a final list of 374 pathogenic deep-intronic SAVs located in 180 genes. Among them, 335 SAVs (89.6%) were reported to cause pseudoexons and 34 SAVs (9.1%) caused extending exons, while the remaining 5 SAVs (1.3%) were reported to cause both pseudoexons and extending exons (compound type) (Fig. 1c). The majority of the variants caused aberrant splicing by creating novel splice donors (59.4%) or splice acceptors (19.5%) (Fig. 1d, e). The other variants were outside the splicing motif regions and might influence splicing enhancers or silencer elements, leading to aberrant splicing.

Benign deep-intronic variants were also collected from WGS data of the 1000 Genomes Project comprised of 2,504 control individuals. We extracted 17,622,722 variants from chromosomes 1, 3, and 5 to reduce the data size (Fig. 1b). After extracting variants on Mendelian diseases genes, variants with ≧5% allele frequency in the 1000 Genomes Project and gnomAD population were collected to assure the benign nature of assorted variants, and the resulting list consisted of 153,794 benign deep-intronic variants. The entire dataset consisting of these pathogenic and benign variants will be called “the curated dataset” hereafter.

### Seeking effective features through characterization of the curated dataset

We first reviewed the characteristics of the curated dataset using SpliceAI, a state-of-the-art splicing predictor constructed on a deep neural network. We scored the variants in the curated dataset with SpliceAI and evaluated the results with a 0.2 threshold, a high-sensitivity threshold provided by the developers (Fig. 1f) [19]. As a result, 338 pathogenic deep-intronic SAVs (90% of 374 SAVs) were above the threshold. This indicates that there was still a need for improvement in sensitivity for clinical use by decreasing the number of false negatives. On the other hand, 190 benign variants exceeded the threshold, resulting in a precision of 64% (338/528). Increasing precision by reducing the number of false positives is essential for the effective prioritization of candidate causative variants. We looked for additional features to support these mispredictions by SpliceAI. For false negatives, we assumed that the result could be effectively complemented by dividing the causes by splice type (SAVs either in splicing motif regions or on the outside (Fig. 1d)). Fourteen SAVs (4.7%) were false negatives in the splicing motif regions (n= 295, in total) (Fig. 1g). The false negatives were thought to be supported by MaxEntScan because it specializes in evaluating the splicing motif regions on the maximum entropy principle, and its superiority to other splice site motif predictors was previously demonstrated [27]. As expected, MaxEntScan predicted these pathogenic SAVs with higher scores than SpliceAI (0.76 vs 0.07 *median values*). On the other hand, of the 45 SAVs outside the splicing motif region, 18 SAVs (40%) were false positives. The tendency toward lower sensitivity for SAVs outside of splice sites than in the splicing motif region is consistent with a previous report (Fig. S2a) [34]. Although the previous report recommended the use of ESRseq scores [35] to support the evaluation of splicing enhancers and silencers, the combination of ESRseq scores and SpliceAI resulted in lower predictive specificity while the sensitivity was improved. In this study, we used the SpliceAI raw score as an alternative. The SpliceAI raw score is the splice site score for the alternative sequence containing the SAV before the score in the reference sequence is subtracted. The score is called the delta score when the reference sequence is subtracted, which is the default output of SpliceAI. We observed that splice site scores on reference sequences were predicted with higher scores for SAVs outside the splicing motif regions than for those within, which might be due to the presence of pre-existing splicing motifs in these regions (Fig S2c). Consistently, for the SAVs within splicing motif regions with ≤0.2 SpliceAI delta scores, the SpliceAI raw scores were higher than the delta scores (0.18 vs 0.06 *median values*). This tendency was not as strong for the SAVs in the splicing motif regions (0.08 vs 0.07 *median values*) (Fig. 1g). This implied that the SpliceAI raw score would improve the predictive sensitivity for SAVs outside the splice sites.

Subsequently, we considered the false-positive results using SpliceAI. We hypothesized that certain predicted variants could cause aberrant splicing, although they had non-deleterious effects on physiological function. However, SpliceAI does not evaluate the deleterious effect of the splicing event because SpliceAI is not trained in pathogenic splicing events. Therefore, a more specific prediction was achieved by combining a deleterious prediction with a human splicing constraint metric from ConSplice, which models mutational constraints on splice-altering variants within the human population. The effectiveness of this approach in predicting deleterious splicing was demonstrated in the ConSpliceML predictor [20]. Likewise, we employed ConSplice for more specialized usage for deep-intronic SAVs. Of the 190 benign variants with ≥0.2 SpliceAI delta score, ConSplice evaluated 153 (81%) variants as being less constrained and having less deleterious effects (<0.2 ConSplice) (Fig. 2h). These observations suggest that the combinatorial use of SpliceAI delta score, SpliceAI raw score, MaxEntScan, and ConSplice would be a better pathogenicity predictor than the sole use of the SpliceAI delta score.

**Fig. 2.**
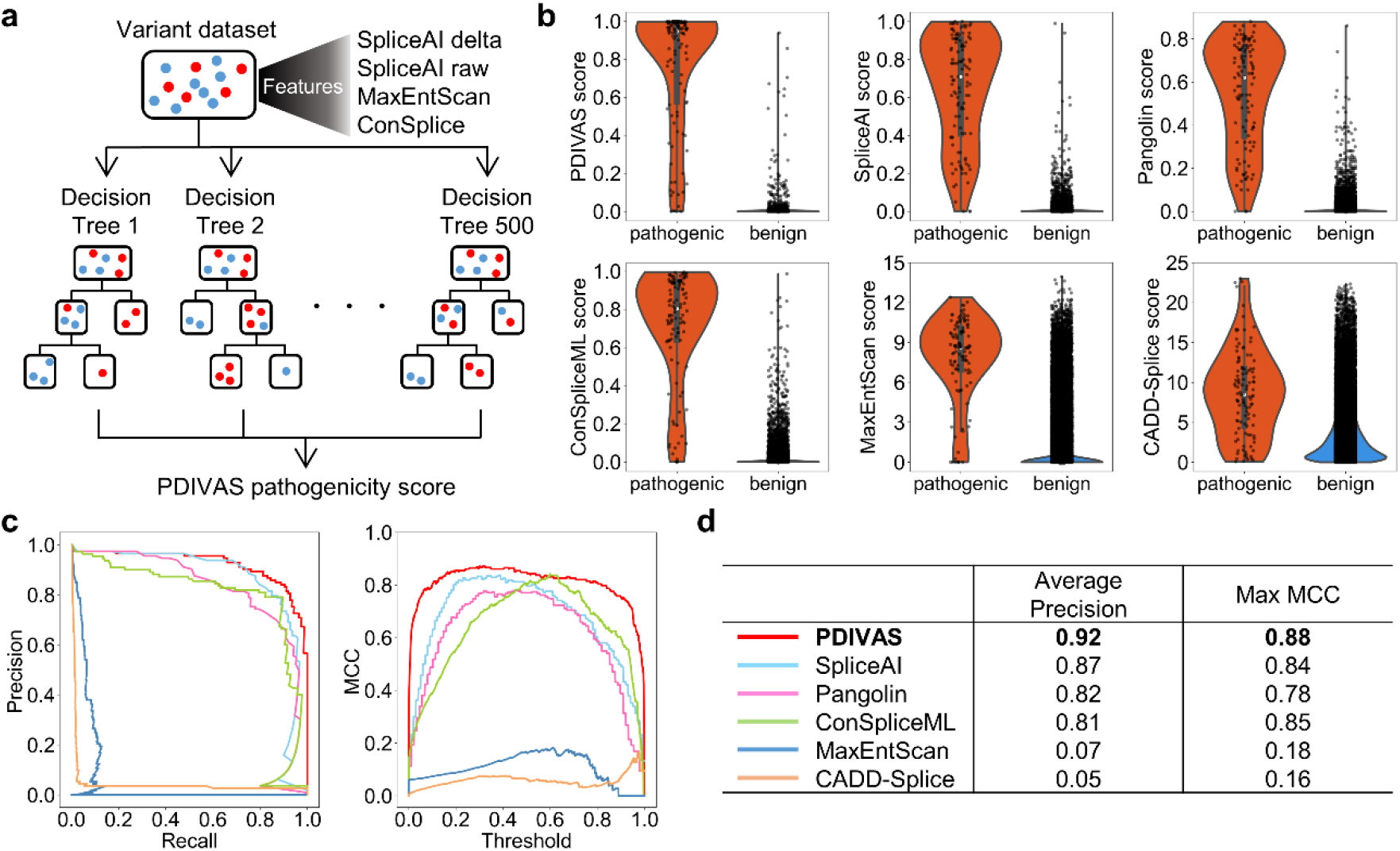
PDIVAS outperforms the existing predictors in classifying pathogenic and benign variants. **a** A graphic of random forest model used in PDIVAS. The model comprised 500 decision trees to classify pathogenic and benign variants (red and blue plots, respectively), combining multiple features from splicing predictors (output-customized SpliceAI and MaxEntScan) and human splicing constraint metrics of ConSplice. **b** Violin plot, box plot, and strip plot indicating the scores of PDIVAS and five previous predictors for the test dataset (pathogenic, n=113; benign, n=46,139). SpliceAI score is represented as the maximum SpliceAI donor/acceptor delta gain scores. Pangolin score is represented as the Pangolin splice gain score. **c** PR and MCC curve analysis for the PDIVAS and five published predictors. Precision, recall, and MCC are calculated at every threshold of their predictors and are depicted as these curves. The MCC values of Pangolin, MaxEntScan, and CADD-Splice are computed on prediction scores scaled from zero to one for the entire curated dataset (min-max normalization). **d** Comparative evaluation of predictive accuracy on average precision and maximum MCC. PR curve, precision-recall curve; MCC, Matthews correlation coefficients

### PDIVAS performs best in predicting pathogenic deep-intronic SAVs

To verify the advantages of using SpliceAI delta gain score (mean and max), SpliceAI raw gain score mean, MaxEntScan, and ConSplice together, we combined these features into one pathogenicity predictor called PDIVAS. PDIVAS is modeled on a random forest classifier where multiple decision trees are built in parallel and each decision tree defines an input deep-intronic variant as pathogenic or benign in a binary manner (one or zero, respectively), referring to these features (Fig. 2a). The PDIVAS calculated the final prediction score as the fraction of trees that classified the variants as pathogenic. Therefore, a higher PDIVAS score indicates a more likely pathogenic splice alteration predicted in the deep-intronic variant, and the score range is between one and zero. To train the random forest model, we randomly split the curated dataset into a training dataset and test datasets. The training dataset represented 70% (261 pathogenic and 107,655 benign variants) of the entire dataset and was used to tune the parameters of the random forest model. The independent test dataset contained the remaining 30% with 113 pathogenic and 46,139 benign variants and was used as a hold-out test dataset to evaluate the final random forest model. The predictive accuracy of the final trained model (PDIVAS) was compared with that of SpliceAI, Pangolin, ConSpliceML, MaxEntScan, and CADD-Splice. Pangolin is a model based on SpliceAI architecture, which was recently proposed. However, its training dataset was augmented by incorporating splice site data detected from four mammalian species into the human dataset. ConSpliceML is a random forest model in which the state-of-the-art splicing predictors of SpliceAI, SQUIRLS, and ConSplice were used as features to classify pathogenic SAV and benign variants. CADD-Splice is an L2-regularized logistic regression model that incorporates various features of conservation scores, transcription factor binding, DNase I hypersensitivity regions, and splicing features driven by the neural network-based splicing predictor of SpliceAI and MMSplice [36].

To compare the predictive accuracy, pathogenic and benign variants were scored with those predictors (Fig. 2b). Subsequently, the precision, recall, and MCC of these predictors at various thresholds were calculated and described as curves (Fig. 2c). A performance comparison of the predictors was performed on the average precision and maximum MCC (Fig. 2d). PDIVAS achieved the highest performance scores (average precision of 0.92 and maximum MCC of 0.88) of the six predictors. We further verified the stable competitiveness of PDIVAS on different training datasets, using five-fold cross-validation on the training dataset. Regardless of the composition of the dataset, PDIVAS was consistently superior to SpliceAI and Pangolin in terms of average precision (0.90 vs. 0.84, 0.77 *median values*) and maximum MCC (0.85 vs. 0.81, 0.76 *median values*) (Fig. S3). These results indicate that PDIVAS outperforms the other state-of-the-art predictors, in predicting pathogenic deep-intronic SAVs.

### Threshold settings for clinical use

Finally, PDIVAS thresholds were set based on the sensitivity of pathogenic SAVs in the PDIVAS test dataset. With the optional threshold, users would be able to change the usage of the PDIVAS, depending on their research objectives and situations (Table 1). For the later performance comparison between predictors, the optional thresholds were also set on SpliceAI and Pangolin on all pathogenic SAVs in the curated dataset.

**Table 1.** Clinically relevant threshold settings on the predictive sensitivity. PDIVAS threshold is set on the test dataset while others’ thresholds are set on the entire dataset. SpliceAI score is represented as the maximum SpliceAI donor/acceptor delta gain scores. Pangolin score is represented as the Pangolin splice gain score.

| Sensitivity (%) | PDIVAS threshold | SpliceAI threshold | Pangolin threshold |
| --- | --- | --- | --- |
| 60 | $\geq 0.899$ | $\geq 0.56$ | $\geq 0.48$ |
| 65 | $\geq 0.832$ | $\geq 0.50$ | $\geq 0.42$ |
| 70 | $\geq 0.763$ | $\geq 0.44$ | $\geq 0.39$ |
| 75 | $\geq 0.575$ | $\geq 0.38$ | $\geq 0.34$ |
| 80 | $\geq 0.501$ | $\geq 0.33$ | $\geq 0.28$ |
| 85 | $\geq 0.340$ | $\geq 0.25$ | $\geq 0.23$ |
| 90 | $\geq 0.151$ | $\geq 0.20$ | $\geq 0.16$ |
| 95 | $\geq 0.082$ | $\geq 0.11$ | $\geq 0.09$ |

### Features in PDIVAS synergistically work to improve predictive accuracy

In this section, we investigate how the five features in PDIVAS were used to improve the predictive accuracy when compared to the sole use of SpliceAI. Firstly, we evaluated the contribution of these features to the extent of which they increased the purity of pathogenic and benign variants in each node of the decision trees (Fig. 3a). This evaluation showed that the most important contributed features of the five were SpliceAI_delta_gain_mean, the mean of the splice acceptor and donor scores. This result is reasonable because the most frequent splice type caused by deep-intronic SAV is a pseudoexon, where both the splice acceptor and donor are newly recognized by spliceosome machinery and both gain scores should be considered (Fig. 1c) (Fig. S4).

**Fig. 3.**
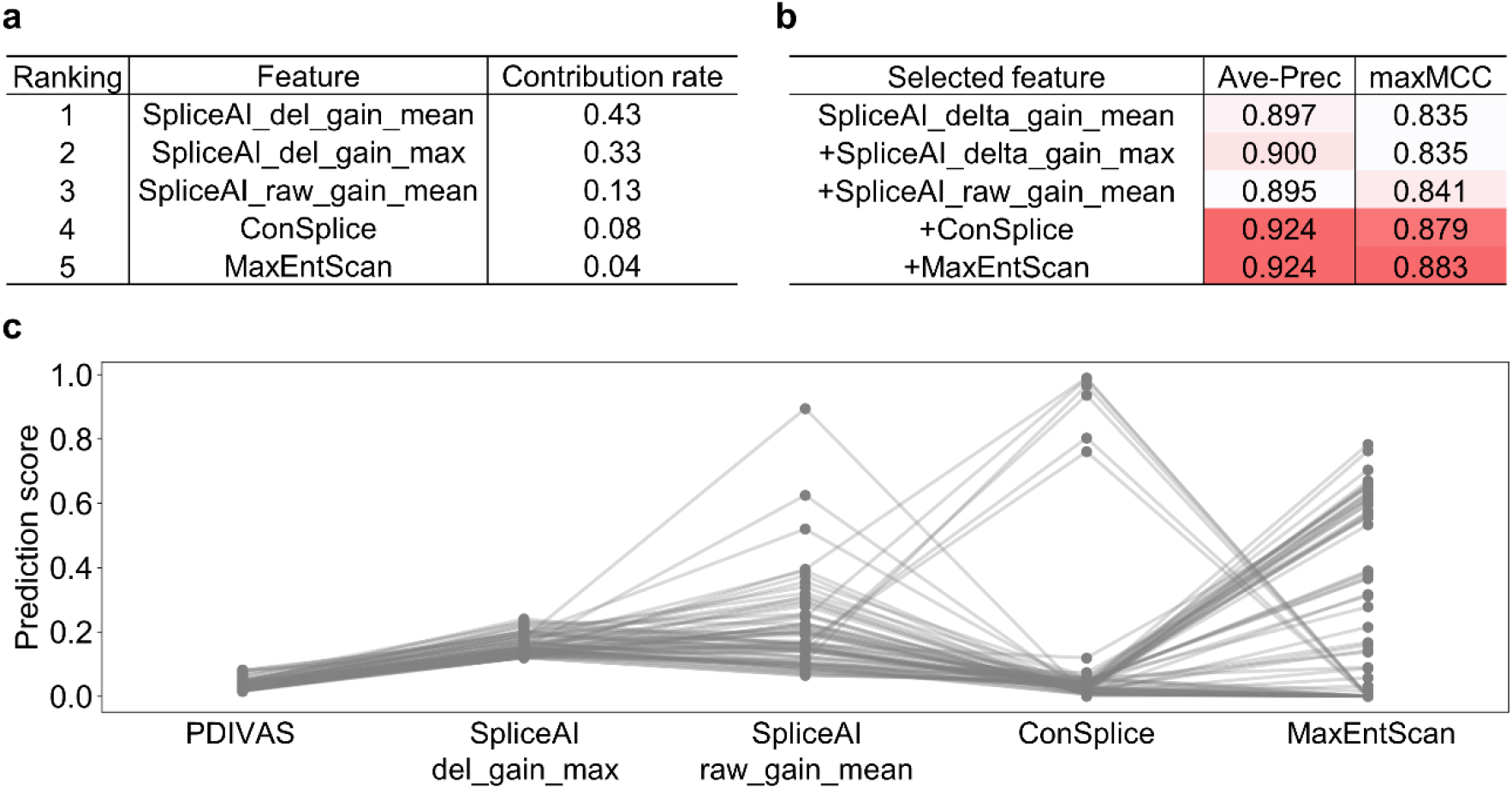
Revealing feature contributions on PDIVAS. **a** The feature contribution rates in the PDIVAS model aree computed as the mean of the accumulation of the impurity decrease within each decision tree. SpliceAI_del_gain_mean indicates the mean values of SpliceAI donor/acceptor delta gain scores. SpliceAI_del_gain_max indicates the maximum of them. SpliceAI_raw_gain_max indicates the maximum of SpliceAI donor/acceptor raw gain scores. **b** The predictive accuracy is evaluated when the random forest model is trained on increased features one by one, starting with only one feature of SpliceAI_del_gain_mean. For each feature subgroup, the same datasets are used to train and evaluate each model. This shows the combinational effect of respective features in improving the average precision (Ave-Prec) or a maximum MCC (maxMCC). **c** Comparison of feature scores of benign variants that are wrongly predicted as pathogenic by SpliceAI (maximums of SpliceAI donor/acceptor delta gain scores), but correctly predicted as benign by PDIVAS. Thresholds are set to 95%-sensitivity mode (Table 1).

Meanwhile, extending exons are caused by the creation of only one of the splice sites, and consistently, one of the SpliceAI gain scores is nearly zero (Fig. S4). Therefore, the SpliceAI_delta_gain_mean might underestimate extending exons. Extending exons were thought to be more appropriately evaluated by the second-most contributed feature, SpliceAI_delta_gain_max only referring to the higher score. The other features SpliceAI_raw_gain_mean, ConSplice, and MaxEntScan also contributed to classifying variants in each node of the decision trees at 13%, 8%, and 4%, respectively.

We further demonstrated the contributions of these features using the feature-cumulative training method (Fig. 3b). This involved training a random forest model with progressively increasing numbers of features, beginning with SpliceAI_delta_gain_mean alone, and evaluating the predictive accuracy of each trained model on the same training/test dataset pair. This analysis showed that each feature of SpliceAI_raw_gain_mean, ConSplice, and MaxEntScan improves either the accuracy metrics of average precision or maximum MCC and has a synergetic effect on PDIVAS. Finally, the feature contributions were studied within the prediction results of benign deep-intronic variants within the test data. When SpliceAI predicted them with 95% sensitivity thresholds, it wrongly evaluated 129 variants as positives. Using PDIVAS with a 95% sensitivity threshold, 73 variants (57%) of the SpliceAI-predicted 129 were correctly evaluated as negatives (Fig. 3c). Overviewing the feature distributions of the 73 variants suggested that PDIVAS removed 67 of them because their genomic regions were in low constraint (<0.2 ConSplice) and the predicted SAVs could be tolerated. The remaining six variants had higher ConSplice scores, but the scores of MaxEntScan were zero and the splice alterations of themselves were not expected to occur. These results demonstrate that PDIVAS succeeded in improving the predictive specificity for benign deep-intronic variants while retaining its high sensitivity for pathogenic deep-intronic SAVs by utilizing these five combined features.

### PDIVAS minimizes candidate variants in WGS samples at clinically relevant thresholds

In the clinical use case of pathogenicity predictors in WGS analysis, the disease-causative variants must be prioritized from the vast number of deep-intronic variants detected in WGS samples. In this study, we analyzed WGS samples of control individuals as a substitute for undiagnosed patients because patients with Mendelian diseases theoretically differ from other individuals by only one to two pathogenic variants. Our analysis focused on the 2,504 control individuals from the 1000 Genomes Project [14]. First, by extracting variants within deep introns in protein-coding genes from each WGS sample, 1,570,571 deep-intronic variants per individual were obtained on a mean value (Fig. 4a, S5a). Assuming the actual genetic diagnosis process, we further extracted variants of 4,429 genes for Mendelian diseases and those with <1% allele frequency in both 1000 Genomes Project and gnomAD populations [18]. This process retained 14,872 variants per individual with possible pathogenicity at a mean value (Fig. S5b, c). Finally, the effects of their variants were predicted using SpliceAI, Pangolin, and PDIVAS. In this part, we chose only the two predictors to compare with because of their higher performance in the analysis of PR and MCC curves and their wider availability for the prediction of genomic insertions and deletions, as well as SNVs. As a result, PDIVAS predicted approximately 3.0-26.8 variant candidates (*mean values*) per individual with thresholds of 70-95% sensitivity while SpliceAI predicted approximately 6.3-67.9 variant candidates, and Pangolin approximately 11.9-136.6 variant candidates (Fig. 4b, Fig. S5d). Even with the most sensitive threshold of 95%, PDIVAS predicted approximately 26.8 candidates, which is approximately 0.4 times and 0.2 times lower than SpliceAI and Pangolin, respectively.

**Fig. 4.**
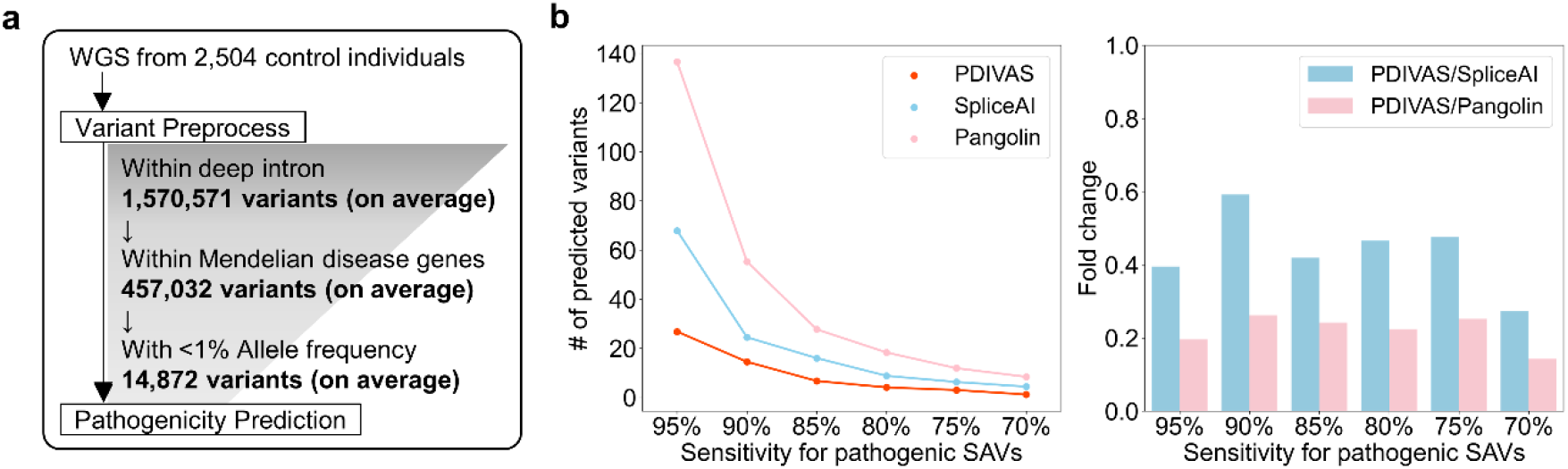
PDIVAS minimizes candidates of causative deep-intronic variants from WGS samples. **a** Workflow of variant filtering for candidates of pathogenic deep-intronic SAVs in 2,504 WGS samples in the 1000 Genomes Project. Each number is shown as the mean value of 2,504 samples. **b** The median numbers of deep-intronic rare variants with PDIVAS, SpliceAI, and Pangolin scores which are higher than the thresholds corresponding to each sensitivity for pathogenic deep-intronic SAVs in the test dataset. To compare the number of predicted variants between PDIVAS, SpliceAI, and Pangolin, fold changes are calculated. For SpliceAI prediction, the maximums of SpliceAI donor/acceptor delta gain scores are used. For Pangolin prediction, gain scores are used.

### PDIVAS performs best in prioritizing causative variants in WGS samples

Lastly, we demonstrated the clinical utility of PDIVAS through the simulation analysis of genetically undiagnosed patients through traditional variant interpretation focusing only on the protein-coding region and exon-intron boundaries. We created virtual patient WGS samples by adding one pathogenic deep-intronic SAV (n=113) in the test dataset to one of the control WGS samples (n=2,504) in the 1000 Genomes Project (Fig. 5a). Using this approach, 282,952 simulated patient WGS samples were obtained.

**Fig. 5.**
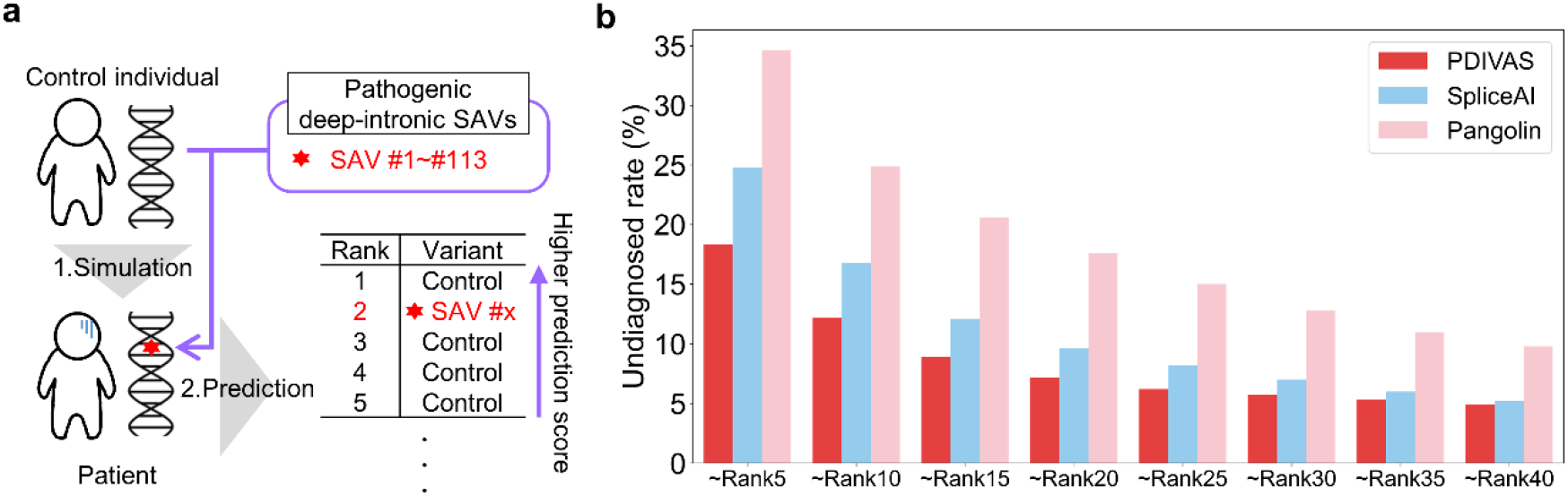
PDIVAS prioritizes causative variants within simulated patient genome sequences. **a** Diagram indicating the method for simulating patient genome sequences and ranking their predicted variants. **b** Undiagnosed rates were calculated as the percentage of simulated patients whose causative variants were not prioritized within the designated ranks, compared to the total number of simulated patients.

This method was originally developed by Danis et al [26]. As shown in Fig. 4a, rare deep-intronic variants were extracted and their pathogenicity was predicted using PDIVAS, SpliceAI, and Pangolin for comparison. The predicted variants were ranked based on their scores, with the assumption that a higher-performance pathogenicity predictor could evaluate pathogenic SAVs with higher scores. Conversely, benign variants with non-deleterious splicing or without splicing outcomes were predicted to have lower scores. As a result, PDIVAS predicted causative SAVs to be significantly closer to the first rank than SpliceAI and Pangolin (Fig. S6). Furthermore, when clinicians manually evaluated up to the 5-40th ranks from the prediction results, more numbers of causative SAVs would be detected through PDIVAS, and the undiagnosed rates of the patients were retained to be 0.3%-6.5% lower than SpliceAI and 4.9%-16.3% lower than Pangolin (Fig. 5b). Even when only the top five were evaluated, 81.7% of the cases would be diagnosed. These results demonstrate that by introducing the diagnostic approach with PDIVAS, clinicians will be able to rapidly find the causative variants because PDIVAS allows them to evaluate a smaller number of candidates and identify the causative variants with minimal effort.

## Discussion

In this work, we have presented PDIVAS, an efficient algorithm to prioritize deep-intronic variants causing aberrant splicing in WGS data. PDIVAS reaches an average precision at 0.92 and a maximum MCC at 0.88, which are the highest among the 6 existing predictors. Even in the WGS analysis, PDIVAS narrows down pathogenic candidate variants into small numbers with 95% sensitivity. Moreover, it ranks most of the causative variants within the top 5 in the simulated patient genome. These results indicate that incorporating PDIVAS into routine analysis pipelines would improve the efficiency of variant interpretation and increase the detection rate of causative variants.

There are three major technical characteristics of PDIVAS. First, the PDIVAS training dataset includes only truly pathogenic SAVs that were checked by us and the curators of HGMD. With only the HGMD curation, the dataset also included variants that are expected to cause splicing, but the events were not experimentally validated although they were given the labels of “Splice” variants. Some actually might not cause aberrant splicing and are not the cause of the diseases. Therefore, we again checked all original reports of pathogenic deep-intronic SAV candidates and extracted experimentally validated ones. Further, we augmented the dataset with that curated by Keegan et al independently of HGMD, where the experimental validation of splice alterations was also strictly checked.

Second, PDIVAS is trained not to classify SAVs and non-SAVs, but to distinguish pathogenic SAVs and benign variants. As some common variants and homozygous variants in general populations are also observed to induce exonization within deep introns, some SAVs do not cause deleterious effects on physiological functions [37]. This indicates that the task of classifying SAVs and non-SAVs is not enough for pathogenicity prediction. Therefore, we incorporated the deleterious effect prediction by ConSplice, as well as splicing features. As shown in Fig. 3c, during PDIVAS prediction, ConSplice helps remove benign SAVs from the SplieAI-predicted SAVs. This prediction of deleterious effect is a technical advantage over SpliceAI and Pangolin, which are not trained on pathogenic splicing events.

Third, PDIVAS is modeled for the specific use of pathogenicity prediction on deep-intronic variants. We divided deep-intronic SAVs into those within the splicing motif regions and those outside of them and observed their SpliceAI-score distributions.

Through detailed observation, we reached the conceptualization of SpliceAI raw scores and MaxEntScan for better sensitivity. To implement their combination, we modeled the random forest optimized to evaluate the pathogenicity of deep-intronic variants. These features were not incorporated into the previous predictor of ConSpliceML.

Determining causative variants is beneficial for patients because it could change their clinical management [38, 39]. The pathogenic pseudoexons created by deep-intronic SAVs can be pharmacologically targeted, as we previously demonstrated in two pathological models: the IVS4+866C>T causative variant of *NEMO* of anhidrotic ectodermal dysplasia with immunodeficiency, and the c.3849+10kbC>T of *CFTR* of cystic fibrosis [40, 41]. Pathogenic pseudoexons often harbor suboptimal splicing sites and are prone to be regulated through alternative splicing factors, and serine/threonine-rich splicing factors (SRSFs) are identified as exon recognition facilitators in the above cases. Thus, the small-molecule inhibitor of CDC-like kinase (CLK) that activates SRSFs through phosphorylation of the RS domain [42], leads to the inhibition of pathogenic pseudoexon recognition and recovery from disease-associated phenotypes in cellular models [40, 41]. The pathogenicity interpretation of the deep-intronic variants with PDIVAS will shed light on previously overlooked pathogenic deep-intronic SAVs in Mendelian diseases, hopefully improving diagnostic rates and the possibility of clinical management.

## Conclusions

Here, we developed a Pathogenicity predictor for Deep-Intronic Variants causing Aberrant Splicing (PDIVAS). The PDIVAS was trained to differentiate between pathogenic and benign deep-intronic variants. The predictive accuracy of PDIVAS was optimized not only by predicting splicing alterations with multiple splicing predictors but also by evaluating the deleterious effect of the predicted splice event with human splicing constraint metrics. By implementing PDIVAS into variant interpretation pipelines, a small number of pathogenic variant candidates were extracted. Efficient variant interpretation by PDIVAS would resolve many genetically undiagnosed cases whose deep-intronic causative variants were previously overlooked. The source code to run PDIVAS and the precomputed PDIVAS scores for all rare deep-intronic SNVs, short insertion, and deletion within Mendelian disease genes are now available at https://github.com/shiro-kur/PDIVAS.

## Data Availability

The PDIVAS source code, command-line interface, and predictions for all rare deep-intronic SNVs, short insertion, and deletion within genes of Mendelian disease are available at https://github.com/shiro-kur/PDIVAS. ConSplice scores and precomputed scores of ConSpliceML are available at https://home.chpc.utah.edu/~u1138933/ConSplice/. Precomputed scores of CADD-Splice are available at https://krishna.gs.washington.edu/download/CADD/v1.6/GRCh37/. The pathogenic splice-altering variants from HGMD were downloaded from the HGMD website http://www.hgmd.cf.ac.uk/ under the HGMD commercial license. Due to HGMD commercial licensing, we are not allowed to share these variants publicly. 1000 Genomes Project variants are publically available at http://hgdownload.cse.ucsc.edu/gbdb/hg19/1000Genomes/phase3/. gnomAD variants are publically available at https://ftp.ensembl.org/pub/data_files/homo_sapiens/GRCh37/variation_genotype/gnomad.genomes.r2.0.1.sites.noVEP.vcf.gz. Gene list from OMIM is available to users from academic institutions and non-profit organizations at https://www.omim.org/downloads. Gene lists from CGD are publically available at https://research.nhgri.nih.gov/CGD/download/.

## Abbreviations

PDIVAS: Pathogenicity predictor for Deep-Intronic Variants causing Aberrant Splicing;
SAV: Splice-altering variant;
HGMD: Human Gene Mutation Database;
SNV: single nucleotide variant;
OMIM: Online Mendelian Inheritance in
Man: CGD Clinical Genomic Database;
VEP: Variant Effect Predictor;
WGS: whole-genome sequencing;
PR: Precision and Recall;
MCC: Matthews correlation coefficient

## Supplementary Information

Additional file 1: Supplementary figures (S1-S6).

## Acknowledgements

We would like to thank members of the M.H. laboratory at Kyoto University for their helpful comments and technical advice, Prof. Zhi-Ming Zheng for reviews on the manuscript, and Editage (www.editage.com) for English language editing. The super-computing resource was provided by Human Genome Center (the Univ. of Tokyo).

## Authors’ contributions

R.K. and K.I. conceived the idea for PDIVAS. R.K. developed the model and performed all analyses. K.I., M.A., T.A., M.Y., K.K., and M.H. provided feedback and suggestions for the PDIVAS modeling. R.K., M.A., and M.H. wrote the manuscript. K.I., M.A., T.A., and M.H. reviewed the manuscript and provided feedback. All authors read and approved the final manuscript.

## Authors’ information

Not applicable

## Funding

This study was supported by JSPS KAKENHI Grant Numbers 22J23899 (to R.K.) and 19K07367 (to M.A.). This study was also supported by AMED under Grant Number JP22gm4010013.

## Availability of data and materials

The PDIVAS source code, command-line interface, and predictions for all rare deep-intronic SNVs, short insertion, and deletion within genes of Mendelian disease are available at https://github.com/shiro-kur/PDIVAS. ConSplice scores and precomputed scores of ConSpliceML [20] are available at https://home.chpc.utah.edu/~u1138933/ConSplice/. Precomputed scores of CADD-Splice [25] are available at https://krishna.gs.washington.edu/download/CADD/v1.6/GRCh37/. The pathogenic splice-altering variants from HGMD [32] were downloaded from the HGMD website http://www.hgmd.cf.ac.uk/ under the HGMD commercial license. Due to HGMD commercial licensing, we are not allowed to share these variants publicly. 1000 Genomes Project variants [14] are publically available at http://hgdownload.cse.ucsc.edu/gbdb/hg19/1000Genomes/phase3/. gnomAD [18] variants are publically available at https://ftp.ensembl.org/pub/data_files/homo_sapiens/GRCh37/variation_genotype/gnomad.genomes.r2.0.1.sites.noVEP.vcf.gz. Gene list from OMIM [30] is available to users from academic institutions and non-profit organizations at https://www.omim.org/downloads. Gene lists from CGD [31] are publically available at https://research.nhgri.nih.gov/CGD/download/.

## Declarations

### Ethics approval and consent to participate

Not applicable.

### Consent for publication

Not applicable.

### Competing interests

The authors declare that they have no competing interests.

## Notes

### Competing Interest Statement

The authors have declared no competing interest.

### Funding Statement

This study was supported by JSPS KAKENHI Grant Numbers 22J23899 and 19K07367. This study was also supported by AMED under Grant Number JP22gm4010013.

### Author Declarations

The study used ONLY openly available human data that were originally located at: http://hgdownload.cse.ucsc.edu/gbdb/hg19/1000Genomes/phase3/. and https://ftp.ensembl.org/pub/data_files/homo_sapiens/GRCh37/variation_genotype/gnomad.genomes.r2.0.1.sites.noVEP.vcf.gz.

### Summary of Updates

Formats of figures were modified (no change in contents) and more detailed descriptions were added to captions.

