## Additional file 1: Supplementary figures (S1-S6) for "PDIVAS: Pathogenicity predictor for Deep-Intronic Variants causing Aberrant Splicing"

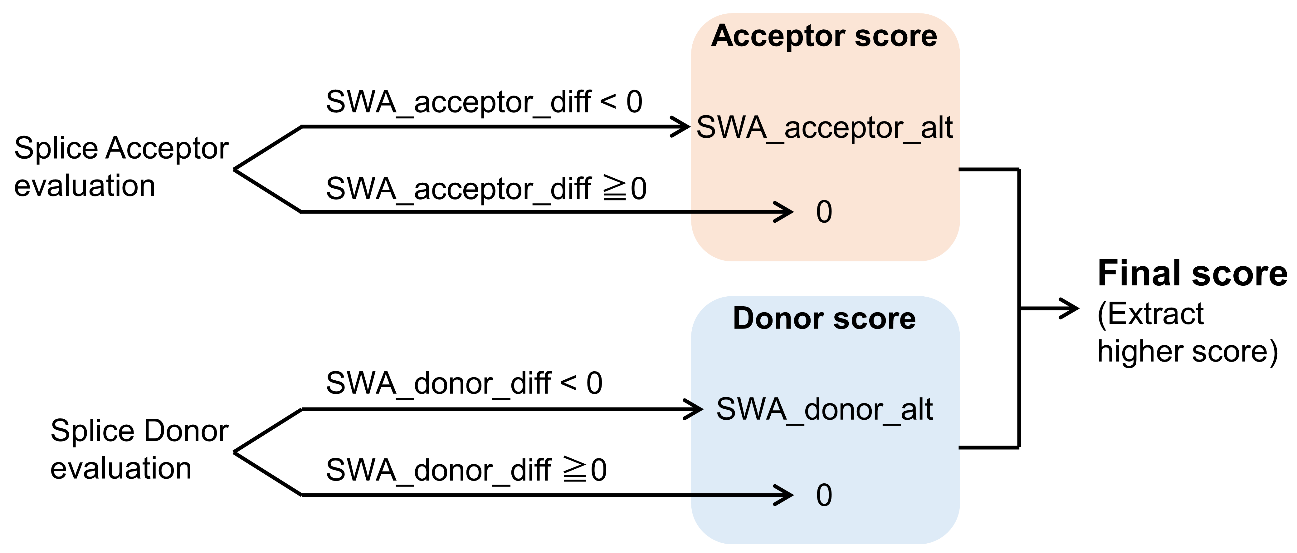

Supplemental Figure 1 Variant-interpreting algorithm of MaxEntScan.

VEP plugged-in MaxEntScan outputs prediction scores for both the splice acceptor and splice donor for each variant. If the splicing motif within the alternate sequence is evaluated more strongly than the reference sequence (SWA_acceptor_diff < 0 / SWA_donor_diff < 0), the raw splice site strengths in alternate sequences (SWA_acceptor_alt, SWA_donor_alt ) are output. Otherwise, zero scores are the output. Finally, a higher score of the splice acceptor and donor evaluation is output as the final score output.

SWA, sliding window algorithm

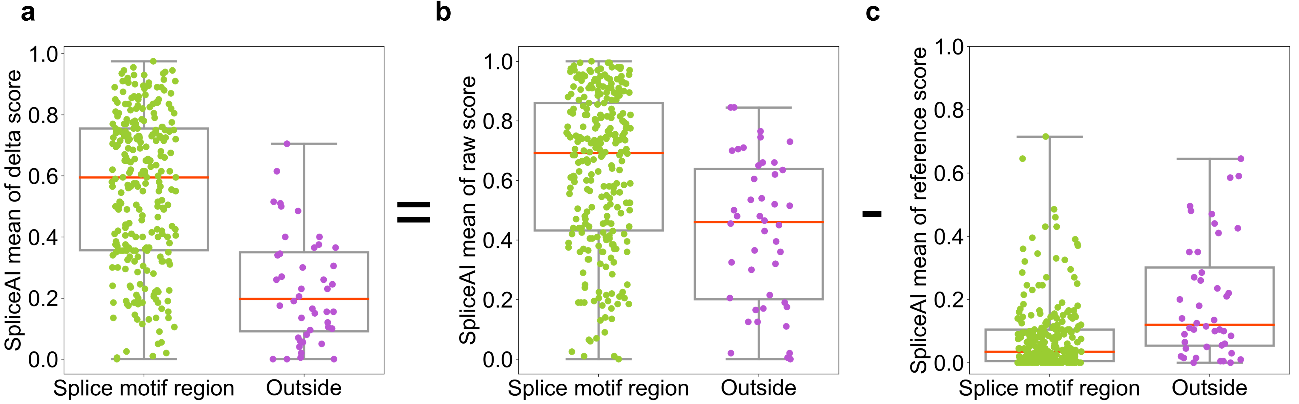

Supplemental Figure 2 SpliceAI raw scores predict SAVs outside of splicing motif regions higher than delta scores.

Box plot and strip plot indicating SpliceAI scores of pathogenic SAVs causing pseudoexons in the PDIVAS entire dataset. The delta scores were calculated by subtracting the reference scores from the raw scores.

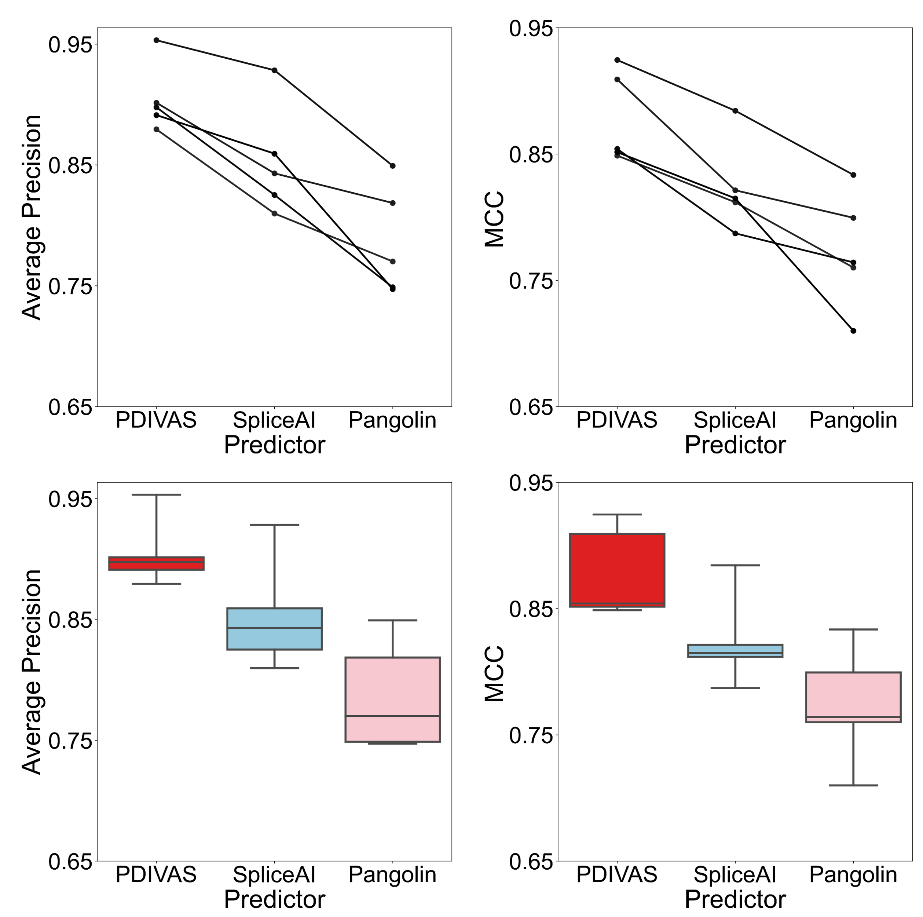

Supplemental Figure 3 PDIVAS has stable competitiveness.

Comparison of predictive accuracy among PDIVAS, SpliceAI, and Pangolin on five-fold cross-validation in the training dataset.

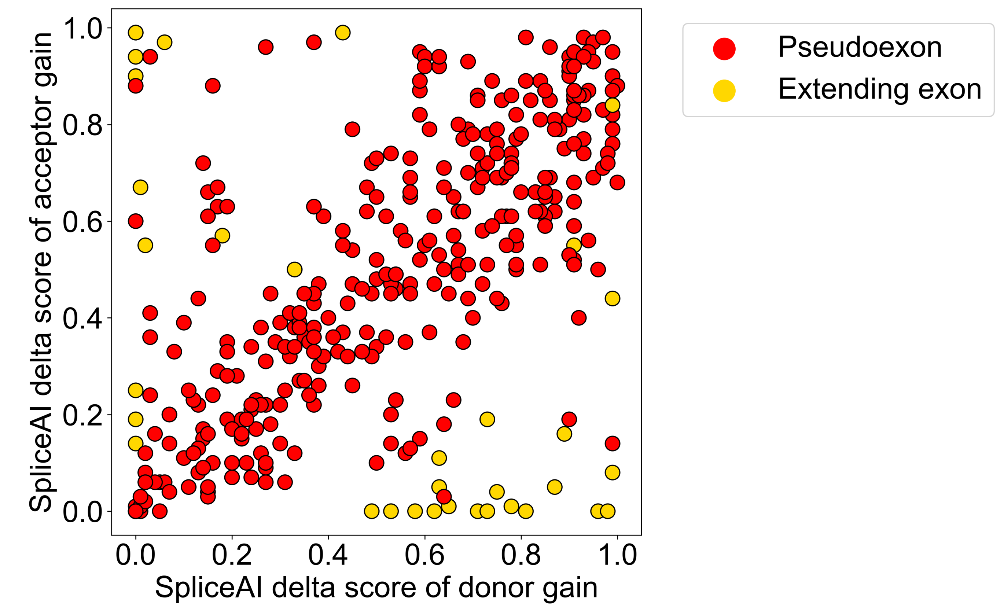

Supplemental Figure 4 One of the SpliceAI scores for pathogenic SAVs causing extended exon often has a score near zero.

The SpliceAI delta scores of acceptor gain and donor gain of pathogenic SAVs causing pseudoexon and extending exon in the entire curated dataset.

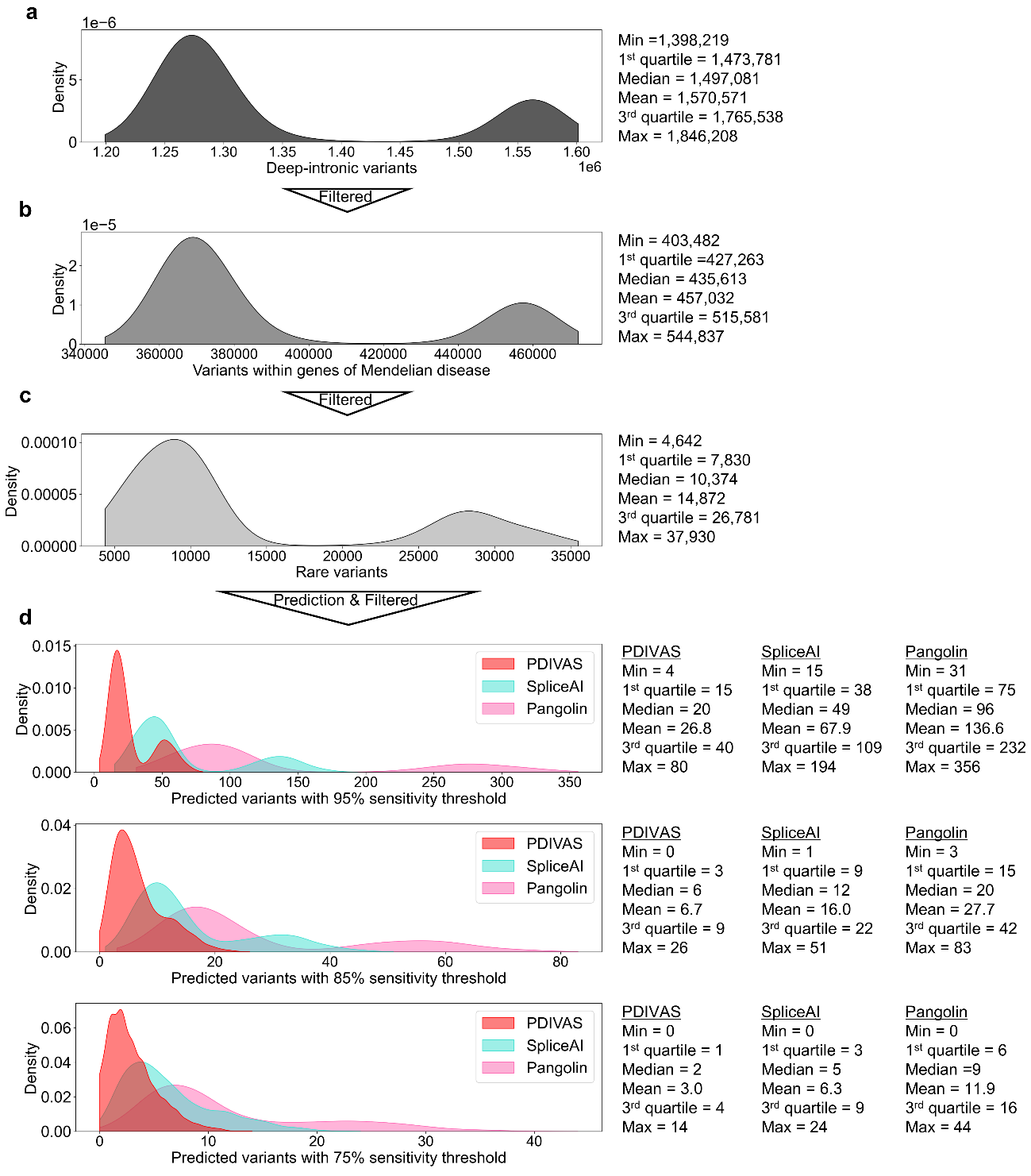

Supplemental Figure 5 PDIVAS extracts the fewest number of candidate variants from control individuals in 1000 Genomes Project (n=2,504).

This result is the raw data of Figure 4.

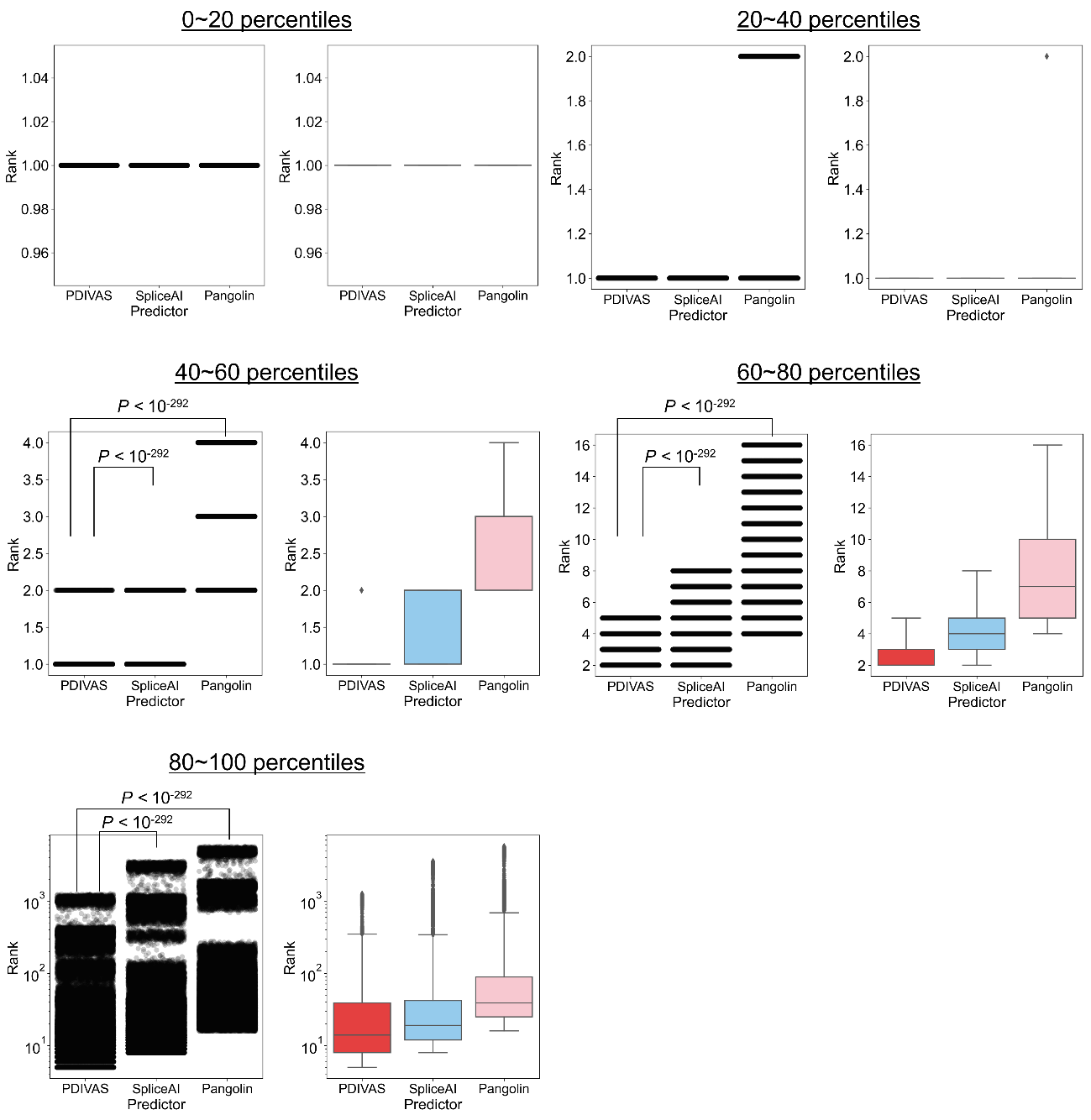

Supplemental Figure 6 PDIVAS predicts causal variants closer to the first rank in simulated patient genome sequences (n=282,952).

This result represents the raw data of Figure 5. The ranks (n=282,952) were sorted from the first to the largest, and separated by percentiles. For each percentile, the ranks are described as strip plots and box plots. The maximum length of the boxplot whiskers is set as ten times the interquartile ranges. Dots outside whiskers are depicted as outliers. P-values were computed using a one-sided Wilcoxon signed-rank test.
